# Validation and optimization of the Japanese version of the Adult Executive Functioning Inventory

**DOI:** 10.1101/2022.11.16.22282379

**Authors:** Toshikazu Kawagoe, Yori Kanekama, Michael J. Rupp

## Abstract

**Background:** Executive function (EF) is an umbrella term used to describe higher-order cognitive processes. Among the test batteries for EF, the Adult Executive Functioning Inventory (ADEXI) is prominent because of several advantages: it is brief, focuses on the core concept of EF, and does not include hard-to-understand general expressions and/or things that are connected but not directly linked to EFs.

**Aims:** To translate the ADEXI into Japanese and reduce the number of items required for optimization. The validation was performed using external self-reporting and laboratory task measurements.

**Methods:** A Japanese version of the ADEXI (J-ADEXI) was created through a regular translation procedure and tested using confirmatory factor analysis (CFA). We also conducted a Mokken scaling analysis (MSA) to reduce the number of J-ADEXI items and correlational analyses with external surveys and laboratory tasks.

**Results:** Although both the original model with 14 items and new the reduced model with 12 items have adequate reliability and validity, the latter was better in terms of model fit. Through discussion, we determined that J-ADEXI includes all 14 items and allows the user to choose a scoring model (i.e., 12 or 14 items model).

**Conclusions:** The J-ADEXI could briefly assess EF with adequate psychometric properties, and this study may also provide clues towards the optimization of the original and/or other language versions of the ADEXI.

## Introduction

Executive function (EF) refers to complex and diverse top-down cognitive functions.^1,2^ Several neuropsychological test batteries targeting EF have been developed, including the Behavioral Assessment of Dysexecutive Syndrome, Cambridge Neuropsychological Test Automated Battery, and Delis–Kaplan Executive Function System. This developmental effort was prompted by the links between EF deficits and psychiatric disorders, including attention deficit hyperactivity disorder, depression, and schizophrenia.^2,3^ However, there is a need for a brief assessment of EF because it cannot take a long time to evaluate clients due to the lack of specialists; for example, in Japan, the position of professional neuropsychologists has yet to be established in clinical settings.^4^ Additionally, in fundamental research, we recently can collect participants’ data online, which enables us to obtain a large sample size despite physical distancing, such as during COVID-19,^5^ wherein researchers may occasionally need to briefly assess EFs as additional variables. In such a situation, the abovementioned batteries are inadequate because considerable time would lead to dropouts.

Holst and Thorell^6^ developed an adult executive functioning inventory (ADEXI). The ADEXI has the advantage of being brief, focusing only on the working memory (WM) and inhibition components of EF. This endeavor is reasonable, especially regarding the limitations of existing questionnaires. First, because attention deficit hyperactivity disorder (ADHD) has the strongest link with executive function,^7^ existing questionnaires frequently include measures to assess ADHD symptoms. Second, because of their aim to assess a wide variety of EFs, they include hard-to-understand general expressions (e.g., the inability to process information quickly or properly) and/or things that are connected but not directly linked to EFs (e.g., easily becoming angry or upset). Third, some of them have too many items (for example, Barkley’s Deficits in Executive Function Scale has 89 items and the Behavior Rating Inventory of Executive Function has 86 items). These constraints were addressed by ADEXI. See the original publication of the ADEXI^6^ and its predecessor, the Childhood Executive Functioning Inventory (CHEXI), for additional information on its development.^8^ Owing to its advantages, the ADEXI has been translated into several languages (see https://chexi.se/), some of which have been validated.^9^ Following this research trend, the current study creats a Japanese version of the ADEXI (J-ADEXI) after following standard translation protocols, including confirmatory factor analysis (CFA). Additionally, this study aimed to optimize the J-ADEXI by reducing the number of items using techniques based on the nonparametric item response theory (IRT), which can be applied in scale development, especially for scale reduction.^10,11^ We tried to validate the full and reduced J-ADEXI not only with an independent questionnaire but also with external behavioral tasks for EF (i.e., flanker task and N-back task) according to traditional standards although it has been argued that the questionnaire survey and experimental test are rarely well correlated, especially in EF research.^12–14^

## Methods

### Preregistration and the circumstances of this study

First, the study was included in a project that aimed to uncover the survey–test association (https://osf.io/fktrp). However, through the translation and validation procedure, it turned out that a single item in the J-ADEXI was not appropriate; therefore, the J-ADEXI required further scrutiny, as reported in the current manuscript. Therefore, we report this questionnaire translation study separately.

### Participants

Two independent samples (Samples 1 and 2) were recruited for this study. Participants were recruited from different data collection companies (iBRIDGE Corporation: https://freeasy24.research-plus.net/ for Sample 1, and CrowdWorks: https://crowdworks.co.jp/en/ for Sample 2). For Sample 1, the sample size was set equal to that of the original ADEXI study,^6^ as registered. However, the results with those data (suppose it as “Sample 0” here; not detailed in this paper) indicated that Item 14 was not appropriate. We speculated that this was due to its low sample size; therefore, we discarded Sample 0 and recruited a greater number of participants for Sample 1, in addition to making minor revisions to Item 14. Therefore, the number of participants in Sample 1 changed from the registered plan. Consequently, our speculation was found to be incorrect, because the results did not change between samples 0 and 1. For Sample 2, the sample size was determined according to the rule of thumb for regression studies as reported elsewhere (see https://osf.io/fktrp for more information about the initial study plan).

Written informed consent was obtained from all the participants. Participants with a previous diagnosis of any mental disorder (e.g., eating disorders, personality disorders, post-traumatic stress disorder, bipolar disorder, mood disorders, anxiety disorders, and schizophrenia) or neurological problems (e.g., seizures, strokes, sleep disorders, and neuropathy) were excluded. Additionally, in both samples, the participants were excluded if they did not respond correctly to the “lure” questions (e.g., “Please check the right-most choices in this item.”) to detect “satisficing” behavior.^15^ The resulting Sample 1 comprised 371 participants (155 women; 52.6 years old on average, ranging from 24 to 75 years), and Sample 2 comprised 327 participants, both of whom were above the minimum requirement for the main analyses.^16^ For Sample 2, another set of participants was excluded because their performance was lower than chance level for each behavioral task and technical problem (two participants). The resulting number of participants analyzed in Sample 2 was 290 (198 women; 39.2 years old on average, ranging from 18 to 75 years).

### Measurements

#### The Japanese version of adult executive functioning inventory (J-ADEXI)

The ADEXI has undergone typical translational procedures, including translation, back-translation, and evaluation (e.g., Tsang et al.^17^). First, the ADEXI was translated by Japanese authors (TK and YK) and back translated by an English native author (MJR) who has a Ph.D. in language education and is naïve to the ADEXI. The back-translated items were checked by Dr. Lisa Thorell, the original ADEXI creator. Based on her evaluations, item(s) were retranslated from the beginning. This procedure was iteratively completed until an agreement was reached between all authors and Dr. Thorell. The J-ADEXI is a 14-item questionnaire with a five-point Likert scale, with higher scores indicating worse EFs. The Japanese, back translated, and original English items are listed in Table S1 in the supplementary material.

#### Effortful control scale (ECS)

To test the construct validity of the J-ADEXI, the Japanese version of the ECS^18^ was administered to Sample 1. The ECS was derived from the Adult Temperament Questionnaire^19^, which was developed as a self-report model of temperament. The ability to use attentional resources and inhibit behavioral reactions, to regulate emotions and related behaviors is referred to as effortful control. The ECS includes inhibitory control (the ability to actively suppress activity), activation control (the ability to initiate behavior even when not motivated), and attentional control (the ability to voluntarily focus or shift attention).^19,20^ As recent empirical findings have reported that effortful control and EF are closely related to an almost unified construct^21^, this can be used as an external rating for concurrent validity. Higher ECS scores indicate greater EFs.

#### Flanker task

A traditional flanker task,^22^ which measures the inhibition of prepotent responses, was administered to Sample 2 via an online platform. The task consisted of presenting a participant with a series of visual stimuli (i.e., arrows pointing right or left in this study) on a computer screen which consisted of a central target stimulus (e.g., an arrow) flanked by distractor stimuli (e.g., four arrows: two for left side of the target and two for right side) that can either be congruent (pointing in the same direction as the target; e.g., “< < < < <“), incongruent (pointing in the opposite direction of the target; e.g., “> > < > >“), or neutral (not providing any direction; e.g., “□ □ < □ □”). As the task was conducted online, the actual stimuli sizes were based on the monitor, which were set as follows: 42 × 50 pixels for the arrow and 17 pixels for the x-axis gap between the arrows. In each trial, following a fixation cross, jittered between 500 and 1500ms, a stimulus array was presented for 1500ms or until a response. The participants were required to respond as quickly and accurately as possible to the direction of the central target stimulus by pressing a left or right arrow on a keyboard. In the experiment, there were a total of 216 trials divided into 3 blocks. In each block, trials were randomly presented under three different conditions: congruent, incongruent, or neutral. Accuracy and reaction time (RT) were measured for this task.

#### N-back task

The N-back task^23^ was used as another behavioral measure for Sample 2. This task involves the presentation of a series of visual stimuli (single capital letters consisting of consonants) and requires the participant to continuously monitor and respond to whether the current stimulus matches the stimulus presented N trials back, where N is a predetermined number (N = 3 was adopted in this study). Regardless of its validity and reliability,^24^ the N-back task is widely used to assess WM function,^25^ because it is considered to have face validity as a WM task.^26^ Participants were required to indicate that the current single letter presented matches the one presented the 3-trials back by pressing a “M” key during the stimulus duration (i.e., 2000ms). Each letter was approximately 80 × 80 pixels in size. The task was divided into three blocks, each containing 50 stimuli, for a total of 150 stimuli with 500ms inter-stimulus-intervals. Only the accuracy was measured.

### Procedure and analysis

Participants answered the questionnaire(s) via a platform provided by each data collection agency. Participants in Sample 1 answered J-ADEXI and ECS, while participants in Sample 2 answered J-ADEXI and engaged in two behavioral tasks: the flanker task and N-back task, which were used to assess inhibition and WM, respectively. The data were obtained and analyzed in the following temporal order. First, the data from Sample 1 were obtained. The J-ADEXI data were analyzed using CFA to assess its construct validity, in which the original ADEXI model was the target to be confirmed (Model 1). Next, to optimize the J-ADEXI by reducing the number of items, we used a technique from nonparametric IRT, the Mokken scaling analysis (MSA^27^). We aimed to reduce the number of items because, as noted in the ‘Participants section,’ the result indicating that one of the items (i.e., Item 14) that appeared inappropriate in Sample 0 was replicated in Sample 1. After reducing the number of items via MSA in Model 2, CFA was conducted again to compare the fit between Models 1 and 2. Construct validity was examined using an external questionnaire (i.e., the ECS) via nonparametric simple and partial correlation analyses in Sample 1. Since the ECS can measure EF as a whole and inhibition, WM, and behavioral initiation as subcomponents, it can be applied to assess the convergent and divergent validities of the J-ADEXI. Additionally, another independent dataset of Sample 2 was obtained for the J-ADEXI and two behavioral measures. We conducted a CFA for model comparison to confirm that Model 2 was better than Model 1 for the J-ADEXI dataset in Sample 2. For simplicity, we used a single standard index for dependent variables in two behavioral tasks: RT cost was calculated for the flanker task (i.e., incongruent RT – neutral RT; the higher, the worse the performance), and d-prime in signal detection theory (the higher, the better the performance) was calculated for the N-back task. Nonparametric correlation analysis was performed to investigate the association between the J-ADEXI and behavioral indices.

Statistical analyses were performed using R (version 4.1.2) running on a PC. CFA using the R lavaan package was conducted to determine whether the original ADEXI factor structure, which is a simple two-factor model (Model 1), could be replicated in the JADEXI. For details on the MSA, see Supplementary Material. The authors assert that all procedures contributing to this work complied with the ethical standards of the relevant national and institutional committees on human experimentation and the Helsinki Declaration of 1975, as revised in 2008. All procedures involving human participants were approved by institutional review board of Tokai University (No. 22020).

## Results

### Confirmatory factor analysis with Sample 1

The items of the J-ADEXI are listed in Table S1 with their corresponding back-translated items and original ADEXI items. The value for Cronbach’s alpha and its 95% confidence interval for the survey in Sample 1 was α = 0.91 (0.90–0.92). Initially, the original two-factor structure (Model 1; see Table S1) was tested using data from Sample 1. The goodness of fit and factor loadings are presented in Tables 1 and 2, respectively. Model 1 seemed to be a good fit for Sample 1, almost satisfying the suggested values;^28^ however, as indicated by the factor loading and communality (Table 2), the variance of Item 14 was poorly explained by the latent factors.

**Table 1.** The goodness of fit indices for all combinations of models and samples.

| Sample/model | CFI | TLI | SRMR | RMSEA | AIC |
| --- | --- | --- | --- | --- | --- |
| 1/1 | 0.906 | 0.888 | 0.056 | 0.076 | 12492.1 |
| 1/2 | 0.924 | 0.906 | 0.046 | 0.091 | 10544.9 |
| 2/1 | 0.840 | 0.808 | 0.068 | 0.087 | 12605.9 |
| 2/2 | 0.879 | 0.850 | 0.057 | 0.083 | 10748.2 |

**Table 2.**
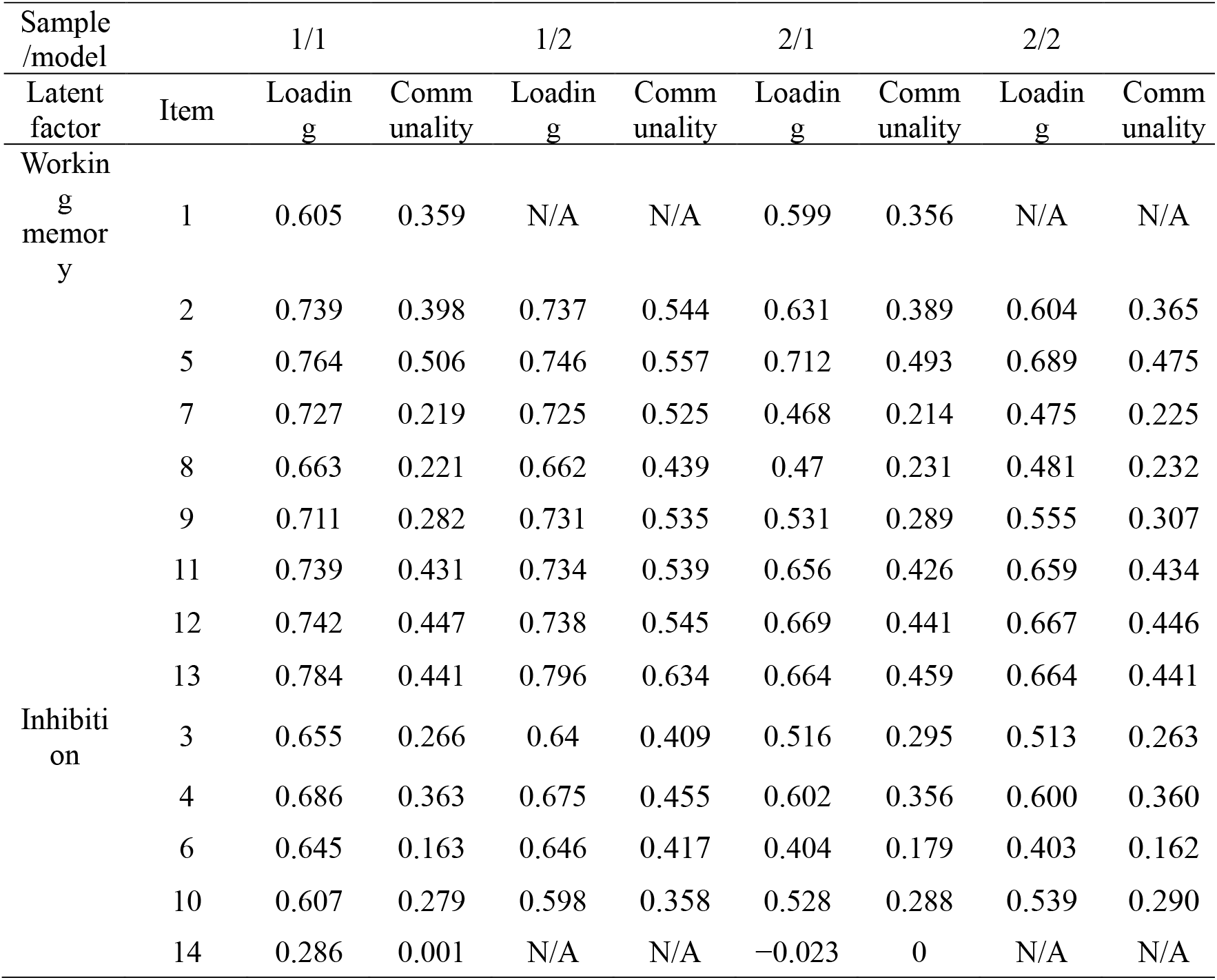
Factor loadings and communality for all combinations of models and samples.

### Item reduction via Mokken scaling analysis with Sample 1

We used Mokken scaling analysis for item reduction of the J-ADEXI, especially the important assumptions (i.e., unidimensionality, monotonicity, and local independence; see Supporting Information for details) that were investigated (see supplementary material for details). For unidimensionality, we can see that all items in each component measure a single latent variable psychologically (i.e., WM or inhibition; Table S1). Mathematically, along with confirming that the two-factor model fits well, we computed Loevinger’s scalability coefficients.^29^ As a rule, scales with *H* < 0.3 are not considered unidimensional or unscalable, scales with 0.3 < H < 0.4 are weak scales, scales with 0.4 < *H* < 0.5 are moderately strong scales, and scales with *H* > 0.5 are strong scales.^27,30^ All items except Item 14 were deemed satisfactory as reasonable scales in Sample 1, as shown in Table S2.

Monotonicity was confirmed for all items except Item 14 based on the scalability coefficients. For additional inspections, the ISRFs and IRFs are shown in Figure S1. Although the IRF indicated no violation of monotonicity in all items, the step 2 ISRF for Item 14 showed a decrease. For local independence, *W* indices were computed (Tables S3 and S4). This analysis revealed that Item 1 was suspected to be locally dependent. After excluding Item 1, no other items were flagged.

After discussion (see Discussion), we decided to exclude Items 1 and 14 from the J-ADEXI. This reduced model (Model 2) has good internal reliability with α = 0.91 (0.91–0.92) as well as the Model 1. A CFA found that all the model fit indices were improved by item reduction (Table 1), and there were no items showing factor loading below 0.5 in Model 2 (Table 2).

### Validation with Sample 1

We then examined convergent validity. In Sample 1, an independent questionnaire for EF (i.e., ECS) was included in the data acquisition, which had three components: inhibitory control, activation control, and attentional control.^19,20^ Theoretically, the subcomponent of inhibition in J-ADEXI is linked to inhibitory control in the ECS, and the WM component in J-ADEXI is linked to attentional control in the ECS. Correlation analyses indicated that all simple correlation coefficients were significant. This is because both questionnaires assessed the same general constructs. Therefore, we calculated the partial correlation coefficients to uncover the specific relationships among the subcomponents of both questionnaires and found that the theoretical hypothesis was supported in both Models 1 and 2 (Table 3).

**Table 3.**
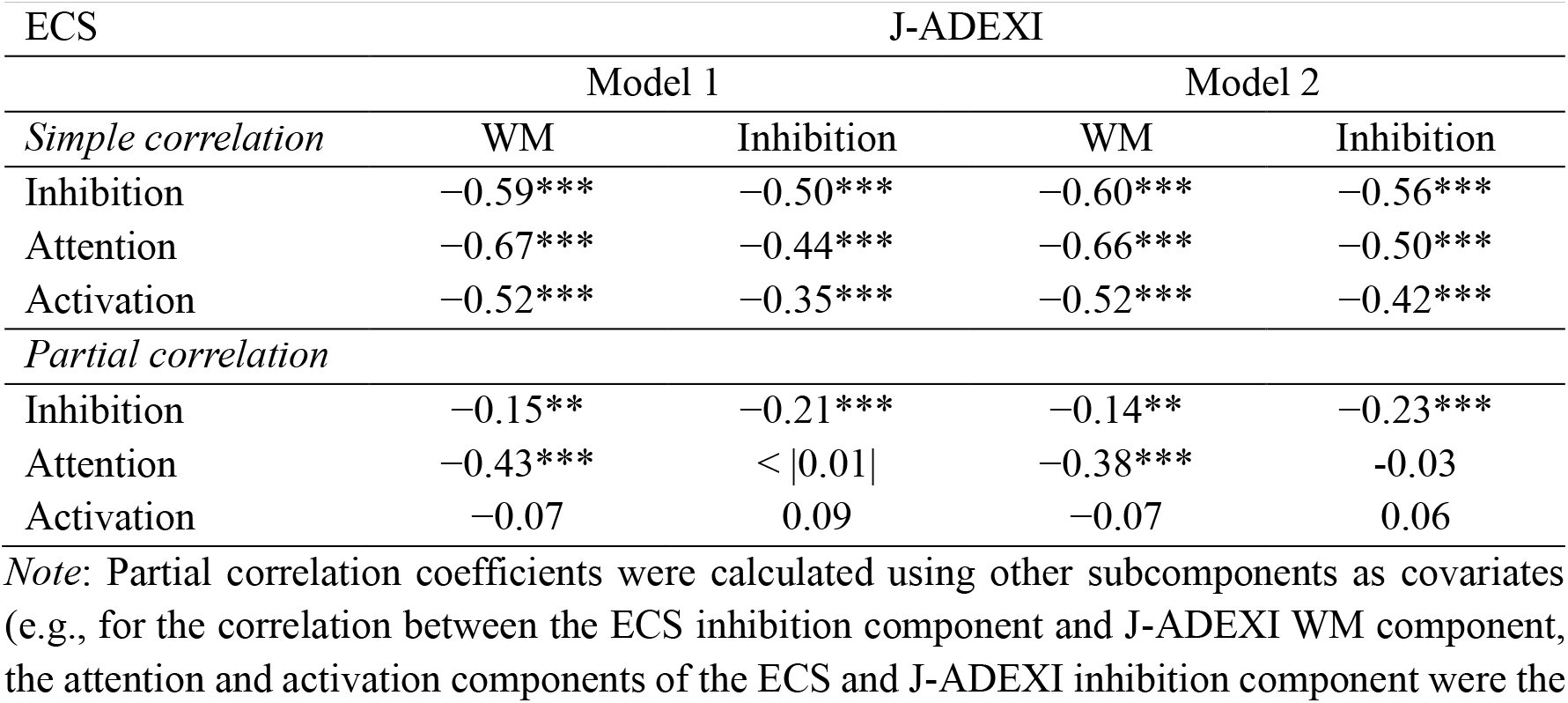
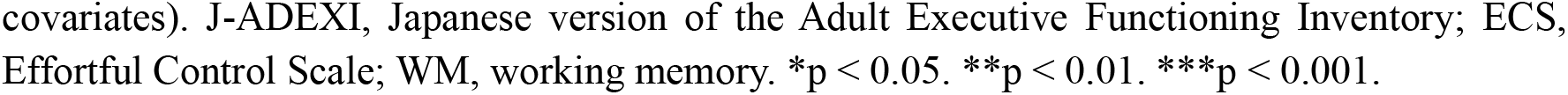
Simple and partial correlation coefficients among both models of J-ADEXI’s two subcomponents and effortful control scale’s three subcomponents in Sample 1.

### Confirmation with Sample 2

After conducting the above series of analyses for Sample 1, data from Sample 2 were obtained, and CFA was applied to both models, which indicated that the results were almost replicated by Sample 2 (Tables 1 and 2). Using Sample 2, we also attempted to confirm the validity of the J-ADEXI with external behavioral indices. Table 4 presents the correlation matrices of the indices. We found a comparable level of correlation between the J-ADEXI and behavioral tasks with the original ADEXI study,^6^ and that there was no difference between Models 1 and 2.

**Table 4.** Correlation coefficients for the association between scores of J-ADEXI and behavioral measures.

| Behaviors | J-ADEXI |  |  |  |  |  |
| --- | --- | --- | --- | --- | --- | --- |
|  | Model 1 |  |  | Model 2 |  |  |
| *Simple correlation* | WM | Inhibition | Full | WM | Inhibition | Full |
| Flanker task | 0.16* | 0.17* | 0.18* | 0.17* | 0.18** | 0.18** |
| N-back task | -0.21** | -0.13 | -0.21** | -0.20** | -0.13 | -0.19** |

*Note:* J-ADEXI, Japanese version of the Adult Executive Functioning Inventory; WM, working memory. \* $p < 0.05$ . \*\* $p < 0.01$ . \*\*\* $p < 0.001$ .

## Discussion

This study aimed to translate the ADEXI into Japanese because it has some advantages compared to other existing questionnaires: ADEXI does not include an item measuring ADHD symptoms, with too general an expression to understand, and does not directly assess EF. Most importantly, it can briefly measure EF by focusing on WM and inhibition, which are the core components of EF.^2,7,31,32^

Although the original model showed a good fit, a single item (i.e., item 14) did not load well. Subsequent MSA supported the CFA results and further indicated that Item 1 was not locally independent. Based on the results, the authors discussed whether these two items should be excluded. First, item 14 can be excluded because the assumption of unidimensionality was not satisfied, in addition to the lower factor loading shown by the CFA. Loevinger’s *H* represents the extent to which items are ordered hierarchically relative to each other, as calculated by the marked value of the items. If an item has a lower value, it means that it has a weaker linear relationship with other test items. The value of item 14 did not exceed the lowest acceptable value of 0.3.^27,30^ Additionally, a question arose regarding the sentence in Item 14: The original item 14 is “People that I meet sometimes seem to think that I am livelier/wilder compared to other people my age.” In the ADEXI, the higher the people rate, the worse their EF. Thus, according to this item, people who think of themselves as liveliers/wilders rather than as people of their age are deemed negative. At least this is unusual in Japan(ese). Therefore, we excluded item 14.

Another item (i.e., Item 1) was also excluded because it was flagged as locally dependent, which means that this item was strongly correlated with other items, even if the latent trait θ was controlled. Although there are several possibilities for the reason for dependency,^33^ in the current case, there might be an unexpected external trait θ′, which defines the item response. We could not find what it was after all, however, through the contemplation of the meaning of the sentence (i.e., “I have difficulty remembering lengthy instructions.”), we claim that it is not an item to assess WM, but rather short-term memory. The item 5 (“When someone asks me to do several things, I sometimes remember only the first or last.”) and item 9 (“I have difficulty planning for an activity.”), which shows the local dependency on item 1, would include short-term memory components rather than Item 1. Therefore, we concluded that items 1 and 14 could be excluded from the J-ADEXI.

The J-ADEXI is more sophisticated than the original ADEXI in terms of the number of items. The exclusion seemed to work well according to the results of the CFA and replication by Sample 2, in which the model fit was better in Model 2 for both samples. The correlation analyses did not show a drastic improvement by item exclusion, but the subcomponents of J-ADEXI maintained a specific relationship with the components of ECS, as theoretically hypothesized: the inhibition component in J-ADEXI correlated with inhibitory control stronger than the other components in ECS, and the WM component did with attentional control in ECS when the other variables were covaried out. For the associations between the J-ADEXI and the two behavioral measures, we found only weak correlations in both models (|r|s < 0.22). However, this would be a replication of the original ADEXI study, in which all the correlations between ADEXI and several EF measures had a small effect size (|r|s < 0.25 in the non-clinical sample).^6^ It should be noted that the literatures have also reported that self-report surveys and laboratory behavioral tests are hardly correlated in EF research; the reasons for this have been suggested as the low reliability of laboratory tasks, that the survey and test could tap different aspect of a psychological construct, or the difference of timescales such as “trait vs state.”^12–14^ Although future studies are warranted because of the low correlations between surveys and tests, we conclude that both scoring models for the J-ADEXI have adequate psychometric properties, similar to the original ADEXI.

We ought to note that all participants were (would-be) normal volunteers although the original study recruited not only normal but also clinical samples.^6^ It is one of the most important characteristics to have the capacity to discriminate whether the rater’s EF is “deteriorated” to the clinical level or not. Although other EF-related tests have been developed and validated only in normal volunteers,^8,34^ future studies must examine the clinical applicability of the J-ADEXI. It can be suggested that J-ADEXI is an additional and supplementary variable because, in addition to its low correlation with tasks, it does not cover every aspect of EFs. ^6^ Further studies are required to determine the significance of these findings. However, the J-ADEXI is a valid and reliable tool for assessing EF, especially WM and inhibition, with only 12 items. Although we found that two of the original items could be excluded, we kept them on the evaluation paper but excluded them only for the calculation step because the translated versions of questionnaires should be equivalent to the original or other language versions (e.g., for cross-cultural studies).^35^ When users need all items, such as for cross-cultural studies, they can calculate the total points, including items 1 and 14, whose availability was shown here by CFA and correlation analysis. It can then be compared with other versions of the ADEXI by excluding two items from the calculation. In conclusion, the J-ADEXI briefly assesses EF with adequate psychometric properties. This study may also provide clues for optimizing the original and/or other language versions of the ADEXI.

## Data Availability

The data underlying the results presented in the study are only available from the corresponding author upon reasonable request because of the data collection agencies' privacy policy.

## Declaration of interests

The authors declare no conflict of interest.

## Funding

This study was supported by grants from the Japan Society for the Promotion of Science (KAKENHI) (grant numbers: 19K14481 and 19H00631).

## Acknowledgements

The authors express great appreciation to Dr. Lisa Thorell for her valuable evaluations and suggestions during the translation procedures.

## Authors’ contributions

TK contributed to the conception and design of the study, data collection, data analysis, interpretation, and drafting of the article. YK and MJR critically revised the translation and manuscript. All authors approved the final version of the manuscript.

## Availability of data and materials

The data, analysis codes that support the findings of this study, and the Japanese scoring sheet are available from the corresponding author, TK, upon reasonable request.

To see the supplementary material, please contact the corresponding author to request the file because the file could not upload to medRxiv because it includes Japanese texts.

## Notes

### Competing Interest Statement

The authors have declared no competing interest.

### Author Declarations

The authors assert that all procedures contributing to this work complied with the ethical standards of the relevant national and institutional committees on human experimentation and the Helsinki Declaration of 1975, as revised in 2008. All procedures involving human participants were approved by institutional review board of Tokai University (No. 22020).

### Summary of Updates

We have included external laboratory (i.e., flanker and N-back) tasks for validation, resulting in indicating the adequate validity of both models in J-ADEXI.

